# Genomics-based timely detection of Dengue Virus type I genotypes I and V in Uruguay

**DOI:** 10.1101/2023.09.05.23295075

**Authors:** Noelia Morel, Marta Giovanetti, Vagner Fonseca, Analía Burgueño, Mauricio Lima, Emerson Castro, Natália R. Guimarães, Felipe C. M. Iani, Victoria Bormida, Maria Noel Cortinas, Viviana Ramas, Leticia Coppola, Ana I. Bento, Alexander Rosewell, Leticia Franco, Jairo Mendez Rico, José Lourenço, Luiz Carlos Junior Alcantara, Hector Chiparelli

## Abstract

We report the first whole-genome sequences of Dengue Virus type I genotypes I and V from Uruguay, including the first cases ever reported in the country. Through timely genomic analysis, identification of these genotypes was possible, aiding in timely public health responses and intervention strategies to mitigate the impact of dengue outbreaks.

**Article Summary Line:** Genomic Analysis for identification of Dengue virus serotype 1 genotypes I and V in Uruguay: an early detection approach

## The Study

Dengue virus (DENV) belonging to the *Flavivivridae* family (genus *Flavivirus*), is a positive-sense, single-stranded RNA virus with a genome spanning approximately 11,000 kb. This pathogen is transmitted by *Aedes aegypti* and *Ae. albopictus* mosquitoes (1), presenting frequent outbreaks with notable public health impact of both mild and severe dengue cases across the Americas, particularly over recent decades (2). DENV’s diversity encompasses four distinct antigenic serotypes (DENV-1 to DENV-4), which are frequently named according to their geographic origins, with a nucleotide variation of approximately 30% (3). Each serotype is additionally subdivided into distinct genotypes (3). Since the beginning of 2023, the Americas have been marked by numerous and substantial dengue epidemics, with nearly three million suspected and confirmed cases documented this year, surpassing the 2.8 million cases reported in 2022 (2). Despite the fact that the Uruguayan Meteorological Institute pinpointed March 2023 (4) as the warmest month in the past 42 years, and with the widespread presence of DENV’s vector mosquitoes throughout the country, Uruguay has so far only experienced intermittent and short-lived outbreaks rather than continuous transmission as observed in neighboring countries. One such outbreak was detected in 2016, involving a total of 20 cases. Subsequently, in 2020, 3 cases were reported. In 2023, a larger, yet restricted outbreak with 35 cases was detected, all attributed to the DENV1 serotype.

Due to the ongoing changes and expansion of transmission in the Americas, including frequent cross-border viral movements (5), continuous and active surveillance remains indispensable to timely detect and manage potential outbreaks in Uruguay. Recognizing this need, we established a collaboration with Montevideo’s Central Public Health Laboratory to conduct a genomics-based study. For this, samples were collected from DENV-positive patients (Ct ≤35) reported and sampled between January 2016 and May 2023 (Table **S1**). This resulted in the generation of the first 24 DENV1 genome sequences of Uruguay (including the first ever reported case: sample ID OR494342) and the identification of co-circulating DENV1 genotypes in the country. Genotype I was traced back to a returning traveler from Asia, marking its first introduction both in the country and the broader region of the Americas. At the same time, genotype V was identified as the dominant circulating in the country. These findings underscore the pivotal significance of a genomics-centered methodology in swiftly detecting emerging viral strains.

Nanopore technology was used to generate genome sequences (DENV1 genotype I n=1, DENV1 genotype V n=23) (Accession numbers: OR494329-OR494352). To put the newly DENV1 genotypes I and V sequence in a global context, we constructed phylogenetic trees including other isolates of the same genotypes. Sequences were aligned using MAFFT (6) and edited using AliView (7). A maximum likelihood phylogeny was reconstructed using IQ-TREE 2 software under the HKY+G4 substitution model (8). Time series data for confirmed, suspected, and probable infections were downloaded from the PAHO website (9). Additionally, climate-driven suitability for DENV (averaged across Uruguay) was estimated using the Index P, as described by Nakase and colleagues (10).

A total number of 24 positive samples tested contained sufficient DNA (2 ng/L) for library preparation. The average cycle threshold (Ct) values for PCR ranged between 15 and 32 (**Table S1**). The sequencing procedure resulted in an average coverage of 94.5%, with a range of 88% to 98.0% (**Table S1**). Cases were reported between January and July, peaking in April, following the natural variation in climate-driven suitability with monthly Pearson’s correlation of 0.92 (**Figure 1A**).

**Figure 1.**
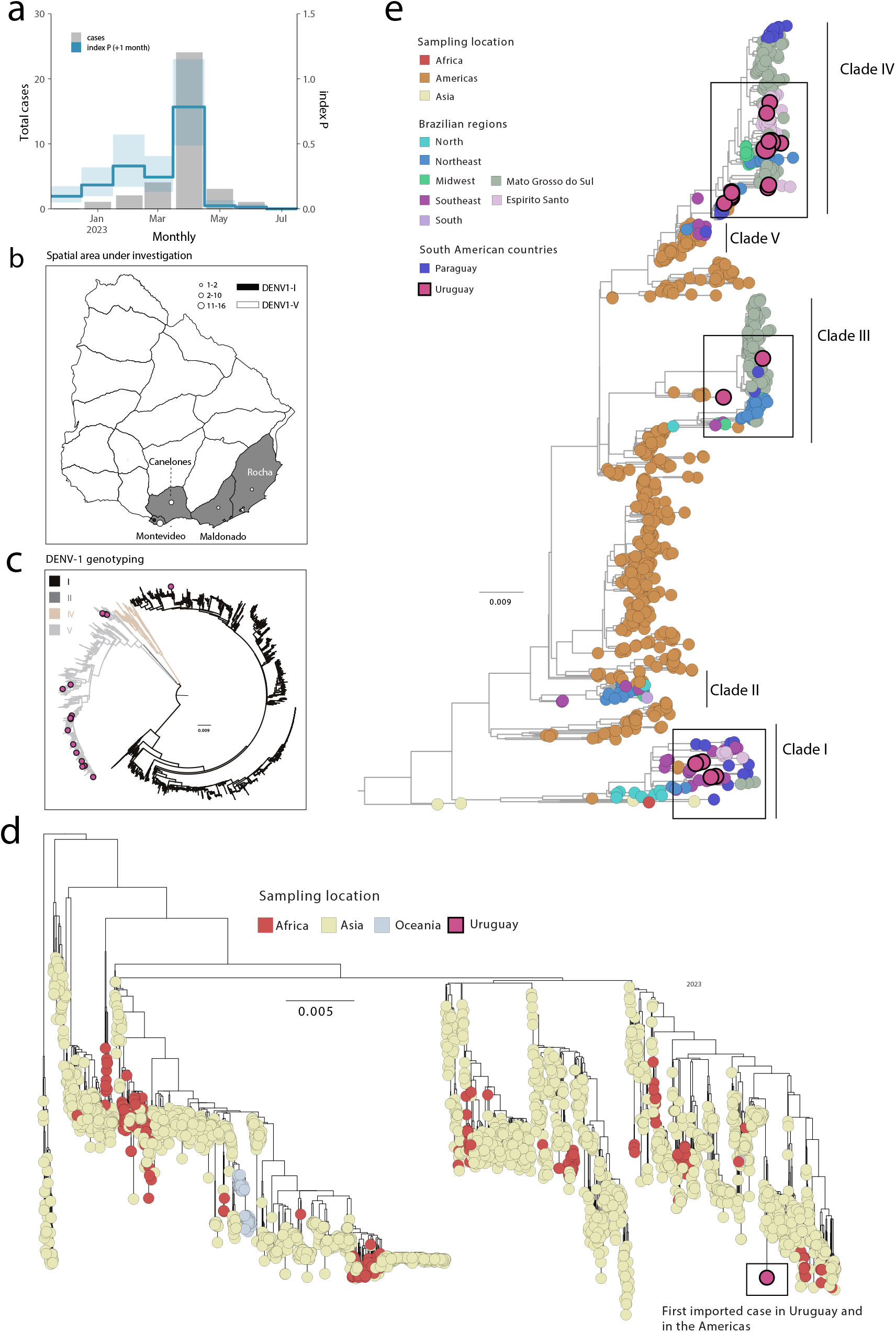
The 2023 DENV1 outbreak in Uruguay. a) Time series of monthly reported DENV cases (grey bars) and index P (climate-driven suitability for DENV transmission: line is the mean, shaded area the 95% interval) in Uruguay in 2023; P is shifted +1 month into the future (the index tends to precede cases, see (9)). b) Map of Uruguay showing the number of DENV1-I and DENV1-V genome sequences obtained per district; c) Maximum likelihood (ML) phylogeny of DENV, including the novel strains described in this study (n=24) in addition to reference sequences representing all genotypes (n=4,141); d) ML phylogeny of DENV1-I genome sequences including the novel strains described in this study (n=1) in addition to reference sequences belonging to the genotype (n=3,004); ML phylogeny of DENV1-V genome sequences including the novel strains described in this study (n=23) in addition to reference sequences belonging to the genotype (n=782).

Four districts were sampled: Canelones (n=4), Maldonado (n=2), Montevideo (17) and Rocha (1) (**Figure 1B**). Genotyping was accomplished utilizing the Dengue virus typing tool (https://www.genomedetective.com/app/typingtool/dengue/) along with preliminary phylogenetic analysis (refer to **Figure 1C**). The results from this analysis confirmed that among the samples, one (n=1) was classified as belonging to Genotype I, while the remaining 23 genomes were categorized as Genotype V. The age of sampled patients ranged from 16 to 66 years, with a median of 39. Among the patients sampled, 70% (n=17) were male (**see Table S1**). Among the cases examined, ten were classified as autochthonous, while the remaining fourteen individuals reported recent travel to various locations, including Argentina (n=1), Asia (n=1), Brazil (n=9), and Paraguay (n=3).

To explore the phylogenetic history of DENV1 genotype I, we combined our newly generated sequence with other DENV1 genotype I genomes available on GenBank (n= 3,004). Our analysis revealed that the sample isolated from a patient with a travel record to Asia grouped with genome sequences from Asia (**Figure 1D**).

To investigate the phylogenetic history of DENV1 genotype V independently, we combined our generated sequences with other DENV1 genotype V genomes available on GenBank (n= 782). Our analysis revealed that the novel isolates belonged to three distinct clades (clade I, III, and IV), which clustered with viral strains isolated from different Brazilian regions and countries in South America, including Paraguay. This result strongly suggested that multiple introductions have taken place within Uruguay (**Figure 1E**) further underscoring the intricate dynamics of viral transmission across geographical boundaries.

## Conclusions

A climate-driven suitability measure is correlated with DENV reporting, highlighting that local climate modulates transmission in Uruguay, thus providing spatio-temporal opportunities for control and mitigation planning. The recent detection of multiple DENV-1 genotypes in Uruguay originating from multiple regions of the globe highlights the necessity of implementing proactive surveillance across the border to prevent the further dissemination of viral variants and mitigate potential future outbreaks.

## Data Availability

Accession numbers: OR494329-OR494352

## Ethics statement

The Pan American Health Organization Ethics Review Committee (PAHOERC) reviewed and approved this project (Ref. No. PAHO-2016-08-0029). The samples used in this research were de-identified residual samples from the routine diagnosis of arboviruses at the Uruguayan public health laboratory, which is part of the Uruguayan Ministry of Health’s public network.

## Biographical sketch

MSc Morel is a distinguished researcher at the Emerging Virus Laboratory, located within the Laboratorio Central de Salud Pública in Montevideo, Uruguay. Her primary focus revolves around molecular diagnosis and sequencing of various critical pathogens, including SARS-CoV-2, arboviruses such as DENV, ZIKV, CHIKV, YFV, and other pathogens that significantly impact Public Health.

## Acknowledgement

This study was supported by the National Institutes of Health USA grant U01 AI151698 for the United World Arbovirus Research Network (UWARN) and the CRP-ICGEB RESEARCH GRANT 2020 Project CRP/BRA20-03, Contract CRP/20/03. M. Giovanetti’s funding is provided by PON “Ricerca e Innovazione” 2014-2020. The authors would also like to acknowledge the Global Consortium to Identify and Control Epidemics – CLIMADE, (T.O., L.C.J.A., E.C.H., J.L., M.G.) (https://climade.health/).

## Conflict of interests

The authors declare that there is no conflict of interests.

## Author contributions

Conception and design: M.G., J.L., L.C.J.A., and H.C.; Investigations: N.M., M.G., V.F., A.B., M.L., E.C., N.R.G., F.C.M.I., V.B., M.N.C., V.R., L.C., L.F., J.L., J.M.R., L.C.J.A., and H.C.; Data Analysis: M.G., V.F. and J.L.; Visualization: M.G. and J.L.; Writing – Original: M.G., J.L.; Revision: N.M., M.G., V.F., A.B., M.L., E.C., N.R.G., F.C.M.I., V.B., M.N.C., V.R., L.C., L.F., J.M.R., J.L., L.C.J.A., and H.C.; Resources: M.G., L.C.J.A., and H.C.

## Notes

### Competing Interest Statement

The authors have declared no competing interest.

### Author Declarations

The Pan American Health Organization Ethics Review Committee (PAHOERC) reviewed and approved this project (Ref. No. PAHO-2016-08-0029). The samples used in this research were de-identified residual samples from the routine diagnosis of arboviruses at the Paraguayan public health laboratory, which is part of the Uruguayan Ministry of Health's public network.

### Summary of Updates

I did a mistake with the author list

## References

1. Adelino, T.É.R., Giovanetti, M., Fonseca, V. et al. Field and classroom initiatives for portable sequence-based monitoring of dengue virus in Brazil. Nat Commun 12, 2296 (2021). 10.1038/s41467-021-22607-0

2. WHO, 2023. https://www.who.int/emergencies/disease-outbreak-news/item/2023-DON475

3. Giovanetti M, Pereira LA, Santiago GA, Fonseca V, Mendoza MPG, de Oliveira C, de Moraes L, Xavier J, Tosta S, Fristch H, de Castro Barbosa E, Rodrigues ES, Figueroa-Romero D, Padilla-Rojas C, Cáceres-Rey O, Mendonça AF, de Bruycker Nogueira F, Venancio da Cunha R, de Filippis AMB, Freitas C, Peterka CRL, de Albuquerque CFC, Franco L, Méndez Rico JA, Muñoz-Jordán JL, Lemes da Silva V, Alcantara LCJ. Emergence of Dengue Virus Serotype 2 Cosmopolitan Genotype, Brazil. Emerg Infect Dis. 2022 Aug;28(8):1725–1727. doi: 10.3201/eid2808.220550. PMID: 35876608; PMCID: PMC9328905.

4. Uruguayan Meteorological Institute, INUMET 2023. https://www.inumet.gub.uy/

5. Vazquez C, Alcantara LCJ, Fonseca V, Lima M, Xavier J, Adelino T, Fritsch H, Castro E, de Oliveira C, Schuab G, Lima ARJ, Villalba S, Gomez de la Fuente A, Rojas A, Cantero C, Fleitas F, Aquino C, Ojeda A, Sequera G, Torales J, Barrios J, Elias MC, Iani FCM, Ortega MJ, Gamarra ML, Montoya R, Rodrigues ES, Kashima S, Sampaio SC, Coluchi N, Leite J, Gresh L, Franco L, Lourenço J, Rico JM, Bispo de Filippis AM, Giovanetti M. Retrospective Spatio-Temporal Dynamics of Dengue Virus 1, 2 and 4 in Paraguay. Viruses. 2023 May 30;15(6):1275. doi: 10.3390/v15061275. PMID: 37376575; PMCID: PMC10304136.

6. Katoh K, Rozewicki J, Yamada KD. MAFFT online service: multiple sequence alignment, interactive sequence choice and visualization. Brief Bioinform. 2019 Jul 19;20(4):1160–1166. doi: 10.1093/bib/bbx108. PMID: 28968734; PMCID: PMC6781576.

7. Larsson A. AliView: a fast and lightweight alignment viewer and editor for large datasets. Bioinformatics. 2014;30:3276–8.

8. Nguyen, L.-T., Schmidt, H.A., von Haeseler, A., Minh, B.Q. IQ-TREE: a fast and effective stochastic algorithm for estimating maximum-likelihood phylogenies. Mol Biol Evol. 2015, 3, 268–74.

9. Pan-American Health Organization, DENV Weekly Report. PAHO. 2023. https://www3.paho.org/data/index.php/en/mnu-topics/indicadores-dengue-en/dengue-regional-en/315-reg-dengue-incidence-en.html

10. Nakase, T., Giovanetti, M., Obolski, U. et al. Global transmission suitability maps for dengue virus transmitted by Aedes aegypti from 1981 to 2019. Sci Data 10, 275 (2023). 10.1038/s41597-023-02170-7

